# Inflammatory biomarkers in pregnant women with COVID-19: a retrospective cohort study

**DOI:** 10.1101/2020.08.27.20183624

**Authors:** Andrea Lombardi, Silvia Duiella, Letizia Li Piani, Ferruccio Ceriotti, Massimo Oggioni, Antonio Muscatello, Alessandra Bandera, Andrea Gori, Enrico Ferrazzi

**Affiliations:** Infectious Diseases Unit, Foundation IRCCS Ca’ Granda Ospedale Maggiore Policlinico, Milano, Italy; Unit of Obstetric, Dept of Woman Child and Neonate, Foundation IRCCS Ca’ Granda Ospedale Maggiore Policlinico, Mangiagalli Centre, University of Milan, Italy; Clinical Laboratory, Foundation IRCCS Ca’ Granda Ospedale Maggiore Policlinico, Milano, Italy; University of Milan, Department of Pathophysiology and Transplantation, Milan, Italy

**Keywords:** pregnancy, coronavirus disease 2019, SARS-CoV-2, inflammatory biomarkers

## Abstract

Coronavirns disease 2019 is a pandemic viral disease affecting also obstetric patients and uncertainties exist about the prognostic role of inflammatory biomarkers and hemocytometry values in patients with this infection. To clarify that, we assessed the values of several inflammatory biomarkers and hemocytometry variables in a cohort of obstetric patients hospitalized with coronavirus disease 2019 and we correlated the values at admission with the need of oxygen supplementation during the hospitalization. Overall, among 27 (61%) pregnant women and 17 (39%) post-partum women, 6 (14%) patients received oxygen supplementation and 2 (4%) required admission to intensive care unit but none died. During hospitalization neutrophils (p=0.002), neutrophils to lymphocytes ratio (p=0.037) and C reactive protein (p<0.001) decreased significantly, whereas lymphocytes (p<0.001) and platelets (p<0.001) increased. Leukocytes and lymphocytes values at admission were correlated with oxygen need, with respectively a 1% and 5% higher risk of oxygen supplementation for each 1,000 cells decrease. Overall, in obstetric patients hospitalized with coronavirus disease 2019, C reactive protein is the inflammatory biomarker that better mirrors the course of the disease whereas D-dimer or ferritin are not reliable predictors of poor outcome. Care to the need of oxygen supplementation should be reserved to patients with reduced leukocytes or lymphocytes values at admission.

## Introduction

The pandemic of coronavirus disease 2019 (COVID-2019), caused by the severe acute respiratory syndrome coronavirus 2 (SARS-CoV-2), is affecting many women during pregnancy and in the postpartum worldwide. According to the available data, the severity of the disease seems comparable between pregnant women and non-pregnant women.^1^ In addition to its impact on the cardio-pulmonary physiology, pregnancy is characterized by several shifts in the woman immunologic profile. Particularly, in preparation of delivery, a pro-inflammatory state occurs with immune cells migrating into the myometrium and high levels of pro-inflammatory cytokines found both in the cervical tissue and in the peripheral blood.^2^ Unfortunately, a peculiar characteristic of COVID-19 is the release of a large amount of inflammatory cytokines, a condition that in severe cases resembles the macrophages activation syndrome.^3^ Some inflammatory biomarkers have been considered as tools to monitor the evolution of COVID-19, namely C reactive protein (CRP), lactate dehydrogenase (LDH), ferritin and D- dimer.^4-6^ Also alterations of the leukocytes count, such as lymphocytopenia or an elevated neutrophils to lymphocytes ratio (NLR), seems correlated with disease severity.^7,8^

Unfortunately, the role of the above-mentioned markers has not been studied during pregnancy and in the postpartum in women infected by SARS-CoV-2. We do not know their evolution during the disease and whether their values can be adopted at admission to guide clinical choices. In this retrospective cohort of obstetric patients admitted with COVID-19 at our tertiary referral centre, we assessed the baseline values and the trend of inflammatory biomarkers and hemocytometry variables and their association with the severity of COVID-19.

## Results

### Demographic and clinical characteristics

We enrolled 44 women. At discharge, 27/44 patients (61%) were still pregnant, whereas 17/44 (39%) delivered during the hospitalization. The median gestational age at admission was 34 weeks and 3 days. Overall, no death occurred in our cohort, 9 (20%) patients required oxygen supplementation, and only 2 (4%) patients were admitted to the intensive care unit (ICU). In both cases for acute pulmonary embolism leading to respiratory insufficiency in patients not receiving prophylaxis with enoxaparin. Pregnant women had lower mean body mass index (p=0.007) and gestational age (p<0.001) compared with those who delivered who instead reached the virologic cure in a longer period (p=0.03) and reported more frequently coryza as symptom (p=0.009). Since March 30, 2020, all patient (26/44, 63%) started at admission prophylactic enoxaparin at the dosage of 100 IU/Kg daily. Immunomodulatory therapy with hydroxychloroquine 200 mg every 12 hours was started in those with lung involvement at chest x-ray (18/44, 41%). Table 1 shows the clinical and demographic characteristics of the patients enrolled.

**Table 1.**
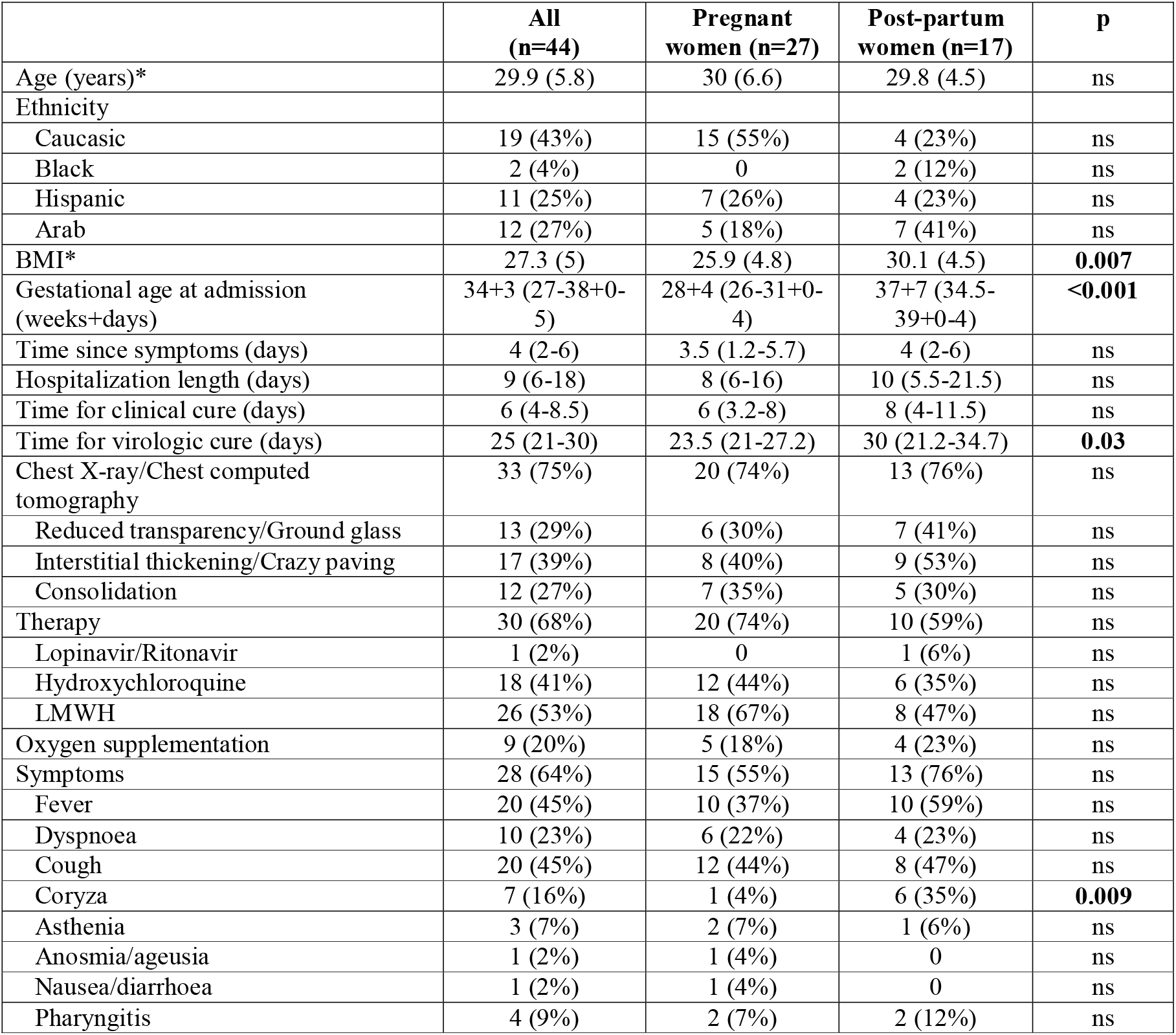
Demographic and clinical characteristics of obstetric patients with COVID-19. Among pregnant women are included those patients who did not deliver during hospitalization, among post-partum women those who did. All data are expressed as median and interquartile range or numbers and percentages except when stated otherwise. *mean (SD) (BMI: body mass index; LMWH: low-molecular-weight heparin; CPAP: continuous positive airways pressure; ICU: intensive care unit)

### Inflammatory biomarkers and white blood cells trend during hospitalization

At admission, the median lymphocytes values were below the lower reference limit (1,200 cells/μL). Instead, the median values of NLR, C reactive protein, D-dimer, procalcitonin and fibrinogen were all above the upper reference limits (3.53, 0.5 mg/dL, 500 μg/L, 0.06 μg/L and 350 mg/dL, respectively). During the observation time neutrophils (p=0.002), NLR (p=0.037) and CRP (p<0.001) values decreased whereas lymphocytes (p<0.001) and platelets (p<0.001) increased. Table 2 summarizes inflammatory biomarkers and white blood cells trend during hospitalization. The results of multiple comparisons tests performed to assess differences among the same variable at different timepoints are shown in figure 1.

**Table 2.** Values (median and interquartile range) of inflammatory biomarkers and hemocytometry variables at different study timepoints. NLR: neutrophils to lymphocytes ratio; LDH: lactate dehydrogenase; AST: aspartate aminotransferase; ALT: alanine aminotransferase.

|  | **Admission** | **48 hours** | **96 hours** | **Discharge** | **p** |
| --- | --- | --- | --- | --- | --- |
| Leukocytes (cells/mL) | 7,790 (5,168-9,405) | 7,790 (5,168-9,405) | 7,150 (5,633-8,640) | 6,735 (5,508-8,563) | ns |
| Neutrophils (cells/mL) | 5,650 (4,595-7,730) | 5,810 (3,110-6,785) | 4,770 (3,540-6,185) | 4,290 (3,425-5,480) | **0.002** |
| Lymphocytes (cells/mL) | 1,180 (1,000-1,830) | 1,675 (1,128-2,008) | 1,545 (1,305-2,058) | 1,675 (1,400-2,145) | **<0.001** |
| Platelets (10<sup>3</sup> cells/mL) | 220 (175-271) | 225 (189-282) | 257 (196-342) | 299 (218-396) | **<0.001** |
| NLR | 4.4 (3.7-5.1) | 3.1 (2.2-5) | 2.9 (2.2-4.4) | 2.8 (2.3-3.6) | **0.037** |
| Ferritin (µg/L) | 38 (27-99) | 56 (34-98) | 67 (28-142) | 51 (24-126) | ns |
| C reactive protein (mg/dL) | 2.7 (1.4-5.4) | 1.9 (1-4.7) | 1.1 (0.3-3.7) | 0.4 (0.2-1.7) | **<0.001** |
| D-dimer (µg/L) | 1,642 (1,298-2,323) | 1,329 (984-1,898) | 1,310 (1,037-1,518) | 1,422 (1,079-1,852) | ns |
| Procalcitonin (µg/L) | 0.08 (0.05-0.1) | 0.08 (0.04-0.1) | 0.06 (0.04-0.12) | 0.05 (0.03-0.09) | ns |
| Fibrinogen (mg/dL) | 450 (399-557) | 462 (421-540) | 485 (397-550) | 430 (365-525) | ns |
| LDH (U/L) | 187 (149-236) | 179 (141-228) | 203 (162-273) | 177 (152-263) | ns |
| AST (U/L) | 43 (24-50) | 32 (13-68) | 55 (45.5-104) | 24 (19-45) | ns |
| ALT (U/L) | 18 (13-22) | 18 (12-25) | 18 (13-28) | 20 (15-35) | ns |

**Figure 1.**
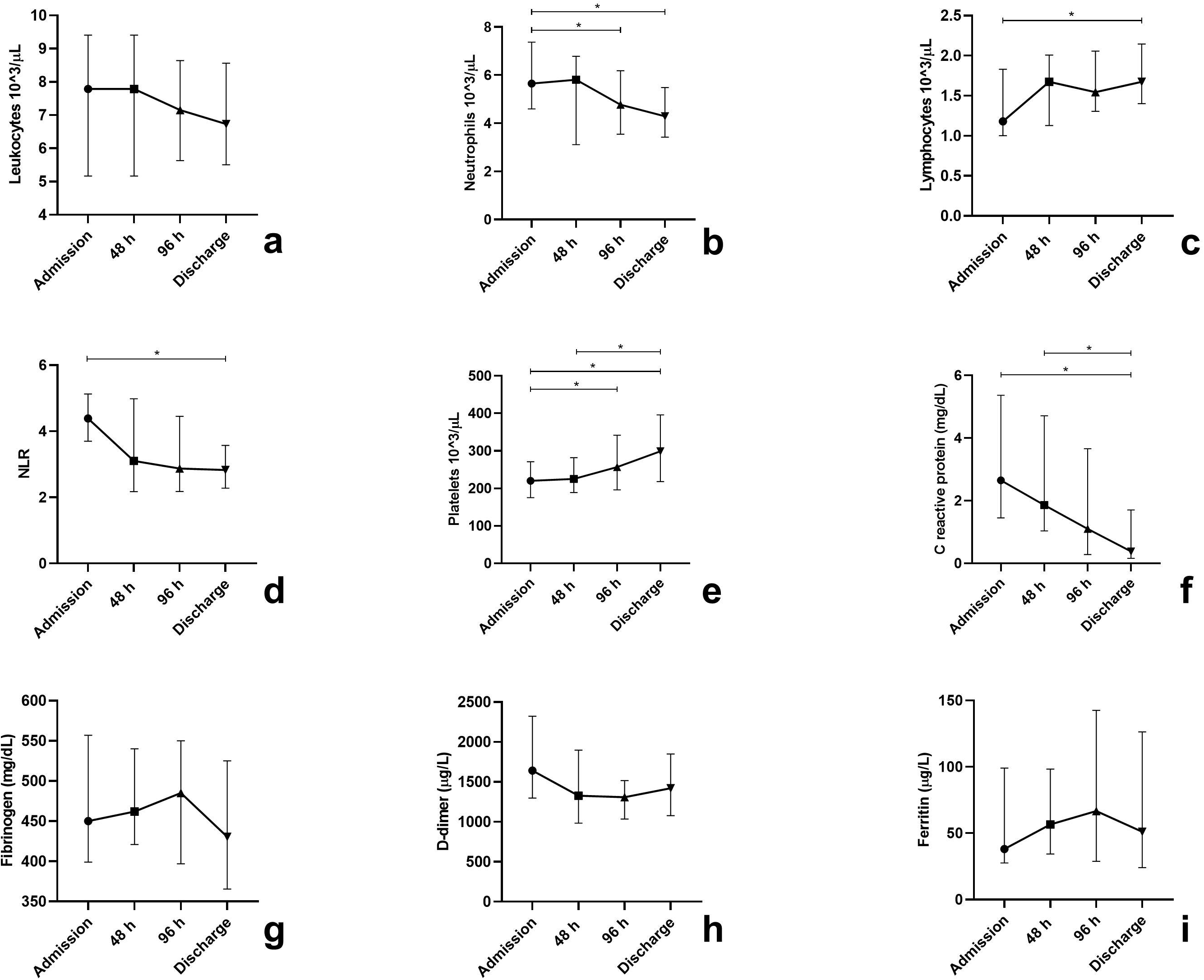
Leukocytes (**a**), neutrophils (**b**), lymphocytes (**c**), neutrophils to lymphocytes (NLR) ratio (**d**), platelets (**e**), C reactive protein (**f**), fibrinogen (**g**), D-dimer (**h**) and ferritin (**i**) assessed at admission, at 48 and 96 hours from admission and at discharge. * denotes statistically significant differences (p<0.005)

### Predictors of oxygen need

The independent effect of different variables at baseline on the likelihood of receiving oxygen supplementation was interrogated by a univariate logistic regression analysis and the results are reported in table 3. The model proved to be significant for leukocytes and lymphocytes and explained 20% and 43% (Nagelkerke R^2^) of the variance for each variable, respectively. Reduced leukocytes and lymphocytes values were associated with an increased likelihood of oxygen supplementation. Two multiple logistic regression models were then built, including in model A leukocytes, CRP and D-dimer and in model B lymphocytes, CRP and D-dimer. Both leukocytes and lymphocytes lost their association with oxygen supplementation in the multivariate model.

**Table 3.** Simple logistic regression and multiple logistic regression models for oxygen supplementation at baseline. In the multivariate logistic regression model A were included leukocytes, CRP and D-dimer, in model B lymphocytes, CRP and D-dimer. NLR: neutrophils to lymphocytes ratio; CRP: C reactive protein; PCT: procalcitonin; CI: confidence interval.

|  | Simple logistic regression |  | Multiple logistic regression |  |  |  |
| --- | --- | --- | --- | --- | --- | --- |
|  |  |  | Model A |  | Model B |  |
|  | Odds ratio<br>(95%CI) | p | Odds ratio<br>(95%CI) | p | Odds ratio<br>(95%CI) | p |
| Leukocytes | 0.999 (0.999-1) | **0.044** | 1 (1-1) | 0.99 |  |  |
| Neutrophils | 0.913 (0.654-1.167) | ns |  |  |  |  |
| Lymphocytes | 0.995 (0.990-1) | **0.037** |  |  | 1 (0.999-1.01) | 0.09 |
| NLR | 1.171 (0.912-1.533) | ns |  |  |  |  |
| CRP | 0.993 (0.890-1.024) | ns | 1.01 (0.950-1.07) | 0.79 | 0.966 (0.888-1.05) | 0.41 |
| PCT | 1.643 (0.905-12.110) | ns |  |  |  |  |
| Fibrinogen | 1.002 (0.996-1.009) | ns |  |  |  |  |
| D-dimer | 0.998 (0.995-1) | ns | 1 (0.99-1.01) | 0.15 | 1 (0.999-1.01) | 0.16 |
| Haemoglobin | 0.910 (0.531-1.561) | ns |  |  |  |  |
| Ferritin | 0.998 (0.975-1.022) | ns |  |  |  |  |

### Discussion

Our findings show how, among the variables assessed in our study, CRP was the only inflammatory biomarkers which varied significantly during the course of COVID-19 in obstetric patients and thus could be employed to monitor the evolution of the disease. Among hemocytometry parameters, neutrophils, lymphocytes, platelets, and neutrophils-to-lymphocytes ratio are those with values which changed significantly. Considering their widespread availability in any hospitalized patient, they also could be employed in assessing disease progression. Interestingly, low leukocytes or lymphocytes values at admission were associated with a higher likelihood of receiving oxygen supplementation during hospitalization, suggesting that additional care should be reserved to patients presenting with reduced values of these blood cells.

Our findings agree with the result of previous studies highlighting how COVID-19 patients are characterized by a relevant systemic inflammation, which acts as a driver of morbidity and mortality. Particularly, lymphocytopenia^8^, a high NLR^9^ and high values of CRP^10^ had all been linked with disease severity or mortality. Our work confirms that these variables show abnormal values also in obstetric patients and mirrors the course of the disease.

Intriguingly, D-dimer values, despite being well above the upper reference limit, were not associated with oxygen need and did not vary significantly during the hospitalization. A mounting amount of evidence is supporting the role of vascular endothelialitis and thrombosis in the pathogenesis of COVID-19^11,12^, and D- dimer values above 1,000 μg/L have been associated with a poor prognosis.^13^ In our cohort D-dimer values were above this limit in all the timepoints. It is possible that the administration of antithrombotic prophylaxis with enoxaparin in more than half of the patients have prevented some additional thromboembolic events. Moreover, it should be recognized that pregnancy is characterized by a physiologic increase of D-dimer values, especially in the third trimester.^14,15^ Therefore, in the specific setting of pregnant women, D-dimer values do not seem a reliable prognostic tool.

To our surprise ferritin values were well above those observed in normal pregnancies,^16^ which is characterized by a physiologic anaemic state and where high levels of ferritin, especially in the third trimester, are associated with negative outcomes (e.g. preterm delivery and gestational diabetes).^17^ At the same time, our values were well below those observed in several cohorts of non-pregnant COVID-19 patients.^18^ For example, Chen and colleagues observed a median ferritin of 337 μg/L among their patients with moderate COVID-19.^6^ Therefore, it is possible to speculate that the inflammatory status caused by SARS-CoV-2 leads to an increase of serum ferritin also in obstetric patients, but this increase is partially concealed by the low levels of this molecule physiologically encountered in this population.

When compared to similar cohorts of obstetric patients with COVID-19, we have a striking similarity in terms of displayed symptoms. Fever, cough and dyspnoea were the three most frequently reported symptoms also in the large British UKOSS (UK Obstetric Surveillance System) cohort and in two Italian cohorts.^19-21^ Interestingly, we have found that the severity of COVID19 in pregnant women or in the post-partum was inferior to that previously reported in the literature. Indeed, we did not observe any death and admission to the ICU was required in only two patients. Instead, Di Mascio and colleagues described in their systematic review a mortality rate of 11.4% and a rate of ICU admission of 31.4%.^22^ Results comparable to ours are observed in several cohorts of obstetric patients with COVID-19 in different regions of the world. Probably these data are more representative of the real-life clinical scenario, being the results provided by Di Mascio *et al*. from a systematic review based on early cases reported in the literature, which usually are those more severe or unusual. New systematic assessments of morbidity and mortality in obstetric COVID-19 patients are needed.

Finally, we observed a longer period of time to reach virologic cure in women who delivered during the hospitalization compared with women who did not. This can be the consequence of a mere methodologic issue. Indeed, per protocol women in the puerperal period were hospitalized for a minimum of 5 days after birth to monitor the new-born. Virologic tests were planned in both groups at 14 days after hospital discharge. Therefore, this difference in time to virologic cure could be only the consequence of a longer permanence in the hospital, even though women in the post-partum did not present a statistically significant longer hospitalization compared to pregnant women. To understand if this difference in viral shedding is real, possibly influenced by different hormonal condition between advanced pregnancy versus earlier phases, a stricter follow-up with repeated molecular assessment of SARS-CoV-2 presence is required.

We recognize that our work has some limitations. First, is a single centre study, and therefore the conclusions can apply only to the specific population afferent to the hospital in which the study was performed. Considering that our hospital was the referring centre for obstetric patients with COVID-19 of a large portion of Lombardy region, the most populous Italian region, and the large variation in ethnicities taken in care, we can suggest that this element did not impact the results. Second, it is a retrospective study, and some analysis (i.d. surveillance nasal swab) were performed for clinical/epidemiologic reasons and not to ascertain biologic differences, as stated above.

Based on the results of our study, a minimum set of analyses composed of hemocytometry plus CRP should be performed in obstetric patients with COVID-19 at admission and during hospitalization, to assess disease severity and follow the evolution of the disease. Specific care to oxygen need should be reserved to those patients presenting with low values of leukocytes and lymphocytes at admission.

## Methods

### Study population

We enrolled a cohort of pregnant women consecutively admitted at the COVID-19 Maternity Hub at the Foundation IRCCS Ca’ Granda Ospedale Maggiore Policlinico in Milan, Italy with a diagnosis of COVID-19 in the period from March 10, 2020 to April 24, 2020. Clinical and demographic data were extracted from the electronic medical records.

The diagnosis of COVID-19 was established by the detection of SARS-CoV-2 in a nasopharyngeal swab. The nasopharyngeal swab was performed either, in all pregnant women admitted for obstetric indication, or in women with at least one of the following signs or symptoms: fever, cough, dyspnoea, asthenia, myalgia, coryza, sore throat, headache, ageusia or dysgeusia, anosmia or parosmia, ocular symptoms, diarrhoea, nausea, and vomit. All SARS-CoV-2 positive cases were admitted in a dedicated COVID-19 ward. Inflammatory biomarkers and hemocytometry variables were assessed at admission, at 48 hours and 96 hours after admission and at discharge. The execution of chest x-ray or chest computed tomography (CT) was reserved to patients with respiratory symptoms, according to clinical judgment. The patients enrolled were subdivided in two groups according delivery during hospitalization or not.

Clinical cure was defined as the absence of fever, a respiratory rate <22 breaths per minutes and peripheral haemoglobin saturation values >95% for at least three days. All patients repeated two nasopharyngeal swabs at an interval of 24 hours, 14 days after discharge or 14 days after clinical cure if still hospitalized for other reasons. If both the swabs resulted negative the patient was considered cured (virologic cure) of the infection and not requiring anymore isolation measures.

### Pro-inflammatory biomarkers assessment and white blood cell profile

Procalcitonin (PCT) and ferritin were measured with electrochemiluminescent immunoassays (ECLIA) on a Roche Cobas e801 instrument (Roche Diagnostics, Monza, Italy). C-reactive protein (CRP) was measured with an immunoturbidimetric method and lactate dehydrogenase (LDH), alanine and aspartate aminotransferase (ALT, AST) with the International Federation of Clinical Chemistry (IFCC) optimized methods on a Roche Cobas c702 instrument (Roche Diagnostics, Monza, Italy). Fibrinogen and D-dimer were measured with ACL Top (Werfen, Milano, Italy). Hemocytometry analysis was performed with Sysmex XN 9000 (Dasit, Cornaredo, Italy).

### SARSCo V-2 detection

Two different methods were used for viral detection. The first one consisted in Seegene Inc reagents (Seoul, Korea), RNA extraction with STARMag Universal Universal Cartridge kit on Nimbus instrument (Hamilton, Agrate Brianza, Italy) and amplification with Allplex® 2019-nCoV assay, while the second employed a GeneFinder® COVID-19 Plus RealAmp Kit (OSANG Healthcare, Anyangcheondong-ro, Dongan-gu, Anyang-si, Gyeonggi-do, Korea) on ELITech InGenius® instrument (Torino, Italy). Both assays identify the virus by multiplex rRT-PCR targeting three viral genes (E, RdRP and N).

### Statistical analysis and ethics

Descriptive statistics were obtained for all the variables reported for this study. Groups were compared with unpaired t test or Mann-Whitney test and Fisher’s exact test. The trend of different variables during hospitalization was assessed with Friedmann test and Dunn’s multiple comparisons test. Univariate logistic regression models were employed to assess correlation between variables at baseline and oxygen supplementation. Multivariate logistic regression models were then built including significant variables at univariate analysis and variables with a biologic correlate. A *p* value <0.05 was deemed statistically significant. All the analysis was performed with SPSS Statistics 23 (IBM Corp, USA). The study was approved by the Institutional Review Board (#339_2020) and conducted according to the Declaration of Helsinki. An informed consent was obtained from all the patients enrolled.

### Data availability

All data will be available on request.

## Data Availability

Alla data will be available on request.

## Authors contributions

AL, EF, AB and AG designed the study. SD and LLP collected the data. AL performed the analysis and wrote the first draft of the manuscript. All the authors revised the final version of the manuscript.

## Acknowledgments

The authors are thankful to Valeria Castelli for the help in data collection.Figure legend

